# Avey: An Accurate AI Algorithm for Self-Diagnosis

**DOI:** 10.1101/2022.03.08.22272076

**Authors:** Mohammad Hammoud, Shahd Douglas, Mohamad Darmach, Sara Alawneh, Swapnendu Sanyal, Youssef Kanbour

## Abstract

**Objectives:** To present our AI-based symptom checker, rigorously measure its accuracy, and compare it against existing popular symptom checkers and seasoned primary care physicians.

**Design:** Vignettes study.

**Setting:** 400 gold-standard primary care vignettes.

**Intervention/Comparator:** We utilized 7 standard accuracy metrics for evaluating the performance of 6 symptom checkers. To this end, we developed and peer-reviewed 400 vignettes, each approved by at least 5 out of 7 independent and experienced general practitioners. To the best of our knowledge, this yielded the largest benchmark vignette suite in the field thus far. To establish a frame of reference and interpret the results of symptom checkers accordingly, we further directly compared the best-performing symptom checker against 3 primary care physicians with an average experience of 16.6 years.

**Primary Outcome Measures:** We thoroughly studied the diagnostic accuracies of symptom checkers and physicians from 7 standard angles, including: (a) *M*1, *M*3, and *M*5 as measures of a symptom checker’s or a physician’s ability to return a vignette’s main diagnosis at the top, among the first 3 diseases, or among the first 5 diseases of their differential diagnosis, respectively (b) *recall* as a measure of the percentage of relevant diseases that are returned in a symptom checker’s or a physician’s differential diagnosis, (c) *precision* as a measure of the percentage of diseases in a symptom checker’s or a physician’s differential diagnosis that are relevant, (d) *F1-measure* as a trade-off measure between recall and precision, and (e) Normalized Discounted Cumulative Gain or *NDCG* as a measure of ranking quality of a symptom checker’s or a physician’s differential diagnosis.

**Results:** Our AI-based symptom checker, namely, Avey significantly outperformed 5 popular symptom checkers, namely, Ada, WebMD, K Health, Buoy, and Babylon by averages of 24.5%, 175.5%, 142.8%, 159.6%, 2968.1% using *M*1; 22.4%, 114.5%, 123.8%, 118.2%, 3392% using *M*3; 18.1%, 79.2%, 116.8%, 125%, 3114.2% using *M*5; 25.2%, 65.6%, 109.4%, 154%, 3545% using recall; 8.7%, 88.9%, 66.4%, 88.9%, 2084% using F1-measure; and 21.2%, 93.4%, 113.3%, 136.4%, 3091.6% using NDCG, respectively. Under precision, Ada outperformed Avey by an average of 0.9%, while Avey surpassed WebMD, K Health, Buoy, and Babylon by averages of 103.2%, 40.9%, 49.6%, and 1148.5%, respectively. To the contrary of symptom checkers, physicians outperformed Avey by averages of 37.1% and 1.2% using precision and F1-measure, while Avey exceeded them by averages of 10.2%, 20.4%, 23.4%, 56.4%, and 25.1% using *M*1, *M*3, *M*5, recall, and NDCG, respectively. To facilitate the reproducibility of our study and support future related studies, we made all our gold-standard vignettes publicly and freely available. Moreover, we posted online all the results of the symptoms checkers and physicians (i.e., 45 sets of experiments) to establish a standard of full transparency and enable verifying and cross validating our results.

**Conclusions:** Avey tremendously outperformed the considered symptom checkers. In addition, it compared favourably to physicians, whereby it underperformed them under some accuracy metrics (e.g., precision and F1-measure), but outperformed them under some others (e.g., *M*1, *M*3, *M*5, recall, and NDCG). We will continue evolving Avey’s AI model. Furthermore, we will study its usability with real patients, examine how they respond to its suggestions, and measure its impact on their subsequent choices for care, among others.

## 1 INTRODUCTION

Digital health has become ubiquitous. Everyday millions of people turn to the Internet for health information and treatment advice [41, 61]. For instance, in Australia, around 80% of people search the Internet for health information, and nearly 40% seek guidance online for self-treatment [13, 27]. In the US, almost two-thirds of adults search the Web for health information and roughly one-third utilize it for *self-diagnosis*, trying to discover by themselves the underlying causes of their health symptoms [36]. A recent study showed that half of the patients investigated their symptoms on search engines before visiting emergency departments [38, 51].

While search engines like Google and Bing are exceptional tools for educating people on almost any matter, they may facilitate misdiagnosis and induce risks stemmed from unrelated health content [36]. This is because Web search entails sifting through an ocean of results, which could emanate from all sorts of sources, and making personal judgements on which data to unveil. Some governments have even launched “Don’t Google It” advertising campaigns to urge their residents to avoid assessing their health using search engines [6, 35]. In fact, search engines are not medical diagnostic tools and laymen are typically not equipped to exploit them for self-diagnosis.

To the contrary of search engines, symptom checkers (referred henceforth to as *checkers*) are patient-facing medical diagnostic tools that mimic clinical reasoning, especially if they use Artificial Intelligence (AI) [2, 27]. They are trained to make medical expert-like judgements on behalf of patients. More precisely, a patient can start a consultation session with a checker via inputting a chief complaint (in terms of one or more symptoms). Afterwards, the checker asks questions to the patient and collects answers from them. Eventually, the checker generates a differential diagnosis (i.e., a ranked list of potential diseases) that explains the causes of the patient’s symptoms.

Checkers are increasingly becoming an integral part of digital health, with more than 15 million users per month [52] that are likely to keep growing [12]. A UK-based study that engaged 1,071 patients found that more than 70% of individuals between the ages of 18 and 39 years would use a checker [16]. A recent study examining a specific checker found that over 80% of patients perceived it to be useful and more than 90% indicated that they would use it again [39]. Various credible healthcare institutions and entities such as the UK National Health Service (NHS) [54] and the government of Australia [43] have officially adopted checkers for self-diagnosis and referrals.

Checkers are inherently scalable (i.e., they can assess millions of people instantly and concurrently) and universally available. Besides, they promise to provide patients with necessary high-quality, evidence-based information [55], reduce unnecessary medical visits [1, 10, 42, 45], alleviate the pressure on healthcare systems [3], improve accessibility to timely diagnosis [1], and guide patients to the most appropriate care pathways [12], to mention just a few.

Nevertheless, the utility and promise of checkers cannot be materialized if they do not prove to be accurate in self-diagnosis [2]. A recent study has showed that most patients (more than 76%) use checkers solely for self-diagnosis [39]. As such, if checkers are not meticulously engineered and rigorously evaluated on their diagnostic capabilities, they may put patients at risk [4, 21, 33]. To this end, this paper focuses on verifying the diagnostic accuracy of checkers due to serving as the underpinning of any aspired benefit.

To begin with, we present ***Avey***, our AI-based checker that was extensively researched, designed, developed, and tested for around 4 years before it was launched. We further propose a thorough scientific methodology that capitalizes on the standard clinical vignette approach for evaluating checkers. Delivering on this methodology, we compiled and peer-reviewed 400 vignettes with 7 external medical doctors using a super-majority voting scheme. To the best of our knowledge, this yielded the largest benchmark vignette suite in the domain. Moreover, we defined and utilized 7 standard accuracy metrics, one of which measures for the first time in the field the ranking qualities of checkers and doctors in generating differential diagnoses.

We leveraged our benchmark vignette suite and accuracy metrics to study the performance of Avey and 5 other major checkers, namely, Ada [23], K Health [26], Buoy [25], Babylon [24], and WebMD [58]. Results show that Avey significantly outperforms the 5 checkers. For instance, Avey outpaced Ada, K Health, Buoy, Babylon, and WebMD by averages of 24.5%, 142.8%, 159.6%, 2968.1%, and 175.5%, respectively in returning the main diagnoses at the top of their differential lists.

In addition, we compared Avey’s performance against 3 highly seasoned primary care physicians with an average experience of 16.6 years. Results reveal that Avey compares favourably to the physicians and even outperforms them with respect to some accuracy metrics, including the ability of ranking diseases correctly within their differential lists and generating the main diagnoses at the top of the lists.

To facilitate the reproducibility of our study and support future related studies, we made our benchmark vignette suite publicly and freely available at [49]. Moreover, we posted all the results of the checkers and physicians at [49] to establish a standard of full transparency and allow for external cross-validation, a step much needed in health informatics [15].

The rest of this paper is organized as follows. We present a high-level overview of Avey’s algorithm in Section 2. Details of our experimentation methodology are given in Section 3 and results are demonstrated in Section 4. We provide a discussion in Section 5 and conclude in Section 6.

## 2 AVEY: A HIGH-LEVEL OVERVIEW

Avey is an interactive medical self-diagnosis system that has been fully researched, designed, and developed in-house. It utilizes an intelligent inference engine with three major components: (1) a diagnostic algorithm, (2) a *finding*^1^ recommendation algorithm, and (3) a ranking mechanism. The inference engine taps into a highly sophisticated probabilistic graphical model, namely, a Bayesian network. Figure 1 demonstrates an actual visualization of Avey’s Bayesian model. The engine’s diagnosis algorithm operationalizes the Bayesian model and generates after every patient’s answer (during a session with Avey) a probability for each modelled disease, conditional on the findings that have been discovered or inferred thus far.

**Figure 1:**
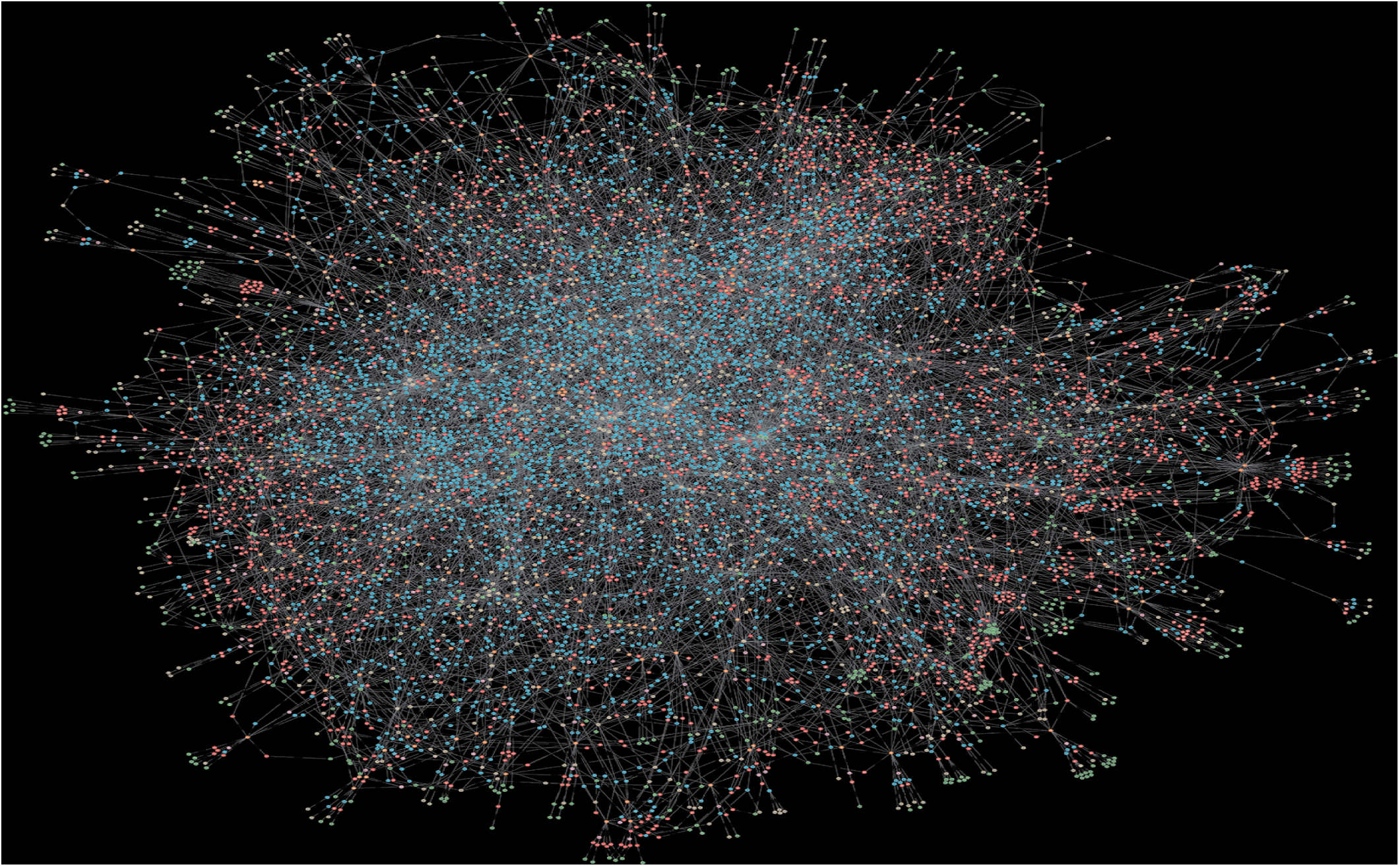
An actual visualization of Avey’s *brain* (i.e., a probabilistic graphical model).

Questions are asked during a patient’s session with Avey via the recommendation algorithm of the inference engine. Specifically, after every answer provided by the patient, the algorithm classifies diseases into three sets, *possible, impossible*, and *unsure*. Subsequently, it predicts the future impact of every relevant finding that has not yet been asked and recommends the one that exhibits the highest impact on the unsure set. The engine asks the recommended finding and continues with the inference process until it converges or hits a maximum number of iterations (or questions). Afterwards, it applies a ranking mechanism that relies on multiple factors to rank all the possible diseases and outputs them as a differential diagnosis to the patient.

## 3 METHODS

### 3.1 Stages

Building on prior related work [12, 22, 27, 36, 52, 53], we adopted a clinical vignette approach to measure the performance of Avey alongside several other checkers. A seminal work at Harvard Medical School has established the value of this approach [22, 52, 53] for testing checkers, especially that it has been also a common method to test physicians on their diagnosis abilities [53].

To this end, we concretely defined our experimentation methodology in terms of 4 stages, namely, *vignette creation, vignette standardization, vignette testing on checkers*, and *vignette testing on doctors*. The 4 stages are demonstrated in Figure 2.

**Figure 2:**
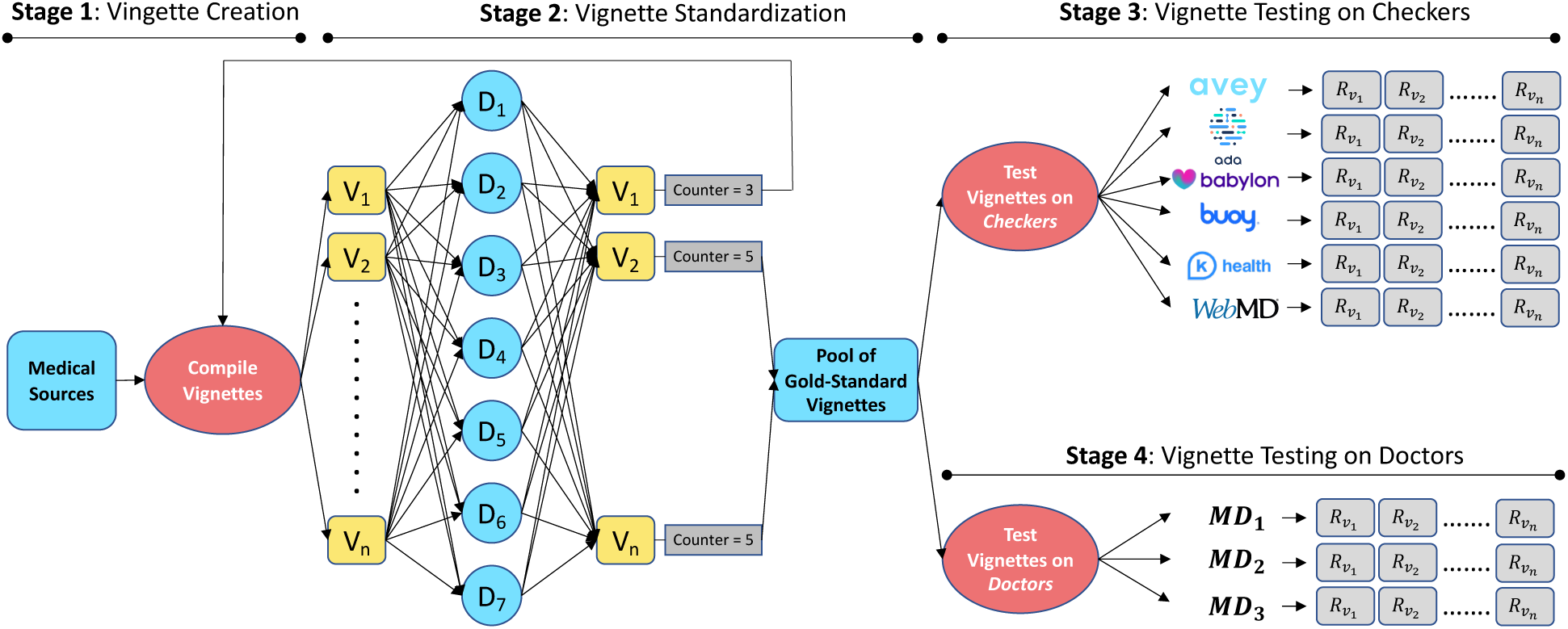
Our 4-stage experimentation methodology (*V*_*i*_ = Vignette *i*, assuming *n* vignettes and 1 ≤ *i* ≤ *n*; *D*_*j*_ = Doctor *j*, assuming 7 doctors and 1 ≤ *j* ≤ 7; *MD*_*k*_ = Medical Doctor *k*, assuming 3 doctors and 1 ≤ *k* ≤ 3; 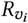 = Result of vignette *v*_*i*_ as generated by a checker or an MD).

In the vignette creation stage, an internal team of medical doctors compiled rigorously a set of vignettes from October 10, 2021 until November 29, 2021. All the vignettes were drawn from reputable medical websites and training material for health care professionals [17, 20, 34, 44, 46, 48, 56, 59]. In addition, our medical team supplemented the vignettes with information that might be “asked” by checkers and physicians in stages 3 and 4. The vignettes involved 14 body systems and encompassed common and less-common conditions relevant to primary care practice (see Table 1). They fairly represent real-world cases in which patients might seek primary care or advice from a physician or a checker.

**Table 1:** The body systems and numbers of common and less-common diseases covered in our benchmark vignette suite.

| Body System | # of Diseases | % of Common Diseases | % of Less Common Diseases |
| --- | --- | --- | --- |
| Hematology | 23 | 8.69 | 91.30 |
| Cardiovascular | 46 | 58.69 | 41.30 |
| Neurology | 22 | 40.90 | 59.09 |
| Endocrine | 20 | 65 | 35 |
| ENT | 23 | 69.56 | 30.43 |
| GI | 44 | 47.72 | 52.27 |
| Obs/Gyn | 54 | 59.25 | 40.74 |
| Infectious | 23 | 26.08 | 73.91 |
| Respiratory | 37 | 70.27 | 29.72 |
| Orthopedics & Rheumatology | 32 | 65.62 | 34.37 |
| Ophthalmology | 18 | 83.33 | 16.66 |
| Dermatology | 12 | 75 | 25 |
| Urology | 14 | 57.14 | 42.85 |
| Nephrology | 32 | 53.12 | 46.87 |

Our medical team constructed each vignette with eight major components: (i) the age and sex of the assumed patient, (ii) a maximum of 3 chief complaints, (iii) the history of the suggested illness associated with details on the chief complaints and other present and relevant findings, (iv) absent findings, including ones that are expected to be solicited by checkers and physicians in stages 3 and 4, (v) basic findings that pertain to physical examinations that can still be exploited by checkers, (vi) past medical and surgical history, (vii) family history, and (viii) the most appropriate main and differential diagnoses.

The output of the vignette creation stage (i.e., stage 1) is a set of vignettes that serves as an input to the vignette standardization stage (i.e., stage 2). Seven medical doctors from 4 specialities, namely, Family Medicine, General Medicine, Emergency Medicine, and Internal Medicine, with an average experience of 8.4 years were recruited from the professional networks of SD, SA, and MD to review the vignettes in this stage. None of these doctors had any involvement with Avey’s project and they were all entirely unaware of it before they were recruited.

We designed and developed a full-fledged web portal to streamline the process of reviewing and standardizing the vignettes. To elaborate, the portal allows our medical team to upload the vignettes to a web page that is shared across the 7 recruited doctors. Each doctor can access the vignettes and review them independently and opaquely (i.e., doctors cannot see the work of each other).

After reviewing a vignette, a doctor can reject or accept it. Upon rejecting a vignette, a doctor can propose changes to improve its quality and/or clarity. Our medical team reviews the suggested changes and makes refinements accordingly, before re-uploading it to the portal for a new round of peer review^2^. Multiple reviewing rounds can occur before a vignette is rendered gold-standard. A vignette becomes gold-standard only if it is accepted by at least 5 out of the 7 (i.e., *super-majority*) external doctors. Once a vignette is standardized, the portal migrates it automatically to stages 3 and 4.

Stage 2 started on October 17, 2021 and ended on December 4, 2021. As an outcome, 400 vignettes were produced and standardized. To the best of our knowledge, this is the largest benchmark vignette suite created to-date to specifically evaluate the performance of checkers. A recent study utilized 200 vignettes and is deemed one of the most comprehensive in the domain thus far [22]. The seminal work of [53] utilized 45 vignettes and many studies followed suit [5, 12, 27, 51]. To allow for external validation and the reproducibility of our results (i.e., the outputs of stages 3 and 4), we made all our vignettes publicly available at [49]. Lastly, we note that none of the 400 vignettes were used in Avey’s development.

The output of stage 2 serves as an input to stage 3, namely, *vignette testing on checkers*. For this sake, we recruited 3 independent primary care physicians from 2 specialities, namely, Family Medicine and General Medicine, with an average experience of 4.2 years from the professional networks of SD and MD. None of these physicians had any involvement with the development of Avey and they were completely unaware of it before they were recruited. Furthermore, two of them were not among the 7 doctors who reviewed the vignettes in stage 2. These doctors were recruited solely to test the gold-standard vignettes on Avey and related checkers.

The approach of having primary care physicians play the role of ‘patients’ in testing checkers has been shown recently to be more reliable than having laypeople doing it [5, 22, 31]. Clearly, laypeople who are not sick and, accordingly, not ‘feeling’ the symptoms or have never felt them will not be able to reliably answer related questions if the answers are not directly contained in the vignettes. In fact, it cannot be guaranteed that checkers will not ask questions that are not contained in the vignettes, even if the vignettes are quite comprehensive. In contrary, physicians can judiciously answer these questions based on the main diagnoses given in the vignettes and figure out whether checkers will be able to converge correctly to these diagnoses.

Besides vignettes, we chose 5 checkers, namely, Ada [23], Babylon [24], Buoy [25], K Health [26], and WebMD [58] to test and compare against Avey. The 5 checkers were selected based on their latest performance results reported in [22], alongside their world-wide popularity with userbases in millions. We tested the vignettes on the most up-to-date versions of these checkers that were available on Google Play, App Store, or websites (e.g., Buoy) between the dates of November 7, 2021 and January 31, 2022.

The 6 checkers (Avey and the 5 competitors) were tested through their normal question-answer flows. As in [22], each of the external physicians in stage 3 randomly pulled vignettes from the gold-standard pool and tested them on each of the 6 checkers (see Figure 2). By the end of stage 3, each physician tested a total of 133 gold-standard vignettes on each checker, except one physician who tested 1 extra vignette to complete the 400 vignettes. Each physician saved a screenshot of each checker’s output for each vignette to allow for results verification, extraction^3^, and analysis. We posted all these screenshots online at [49] to establish a standard of full transparency and allow for external cross-validation and study-replication.

In stage 4, we recruited 3 more independent and experienced primary care physicians with an average experience of 16.6 years from the professional networks of SD, SA, and MD. One of those physicians is a Family Medicine doctor with 30+ years of experience. The other two are also Family Medicine doctors, each with 10+ years of experience. None of these physicians had any involvement with the development of Avey and were completely unaware of it before they were recruited. Furthermore, none of them were among the 7 or 3 doctors of stages 2 or 3, respectively and were only recruited for conducting stage 4.

The solo aim of stage 4 is to compare the accuracy of the winning checker against that of experienced primary care physicians. Hence and akin to [52], we concealed the main and differential diagnoses of the 400 gold-standard vignettes from the 3 recruited doctors and exposed the remaining information through our web portal. The doctors were granted access to the portal and asked to provide their main and differential diagnoses for each vignette without checking any reference, mimicking as closely as possible real-world sessions where they typically diagnose patients on the spot without checking references. As an outcome, each vignette was ‘diagnosed’ by each of the 3 doctors. The results of the doctors were posted online at [49] to allow for external cross-validation.

### 3.2 Accuracy Metrics

To evaluate the performance of checkers and doctors in stages 3 and 4, we utilize 7 standard accuracy metrics. As in [19, 22], for every tested gold-standard vignette, we use the matching-1 (***M1***), matching-3 (***M3***), and matching-5 (***M5***) criteria to measure if a checker or a doctor is able to output the vignette’s main diagnosis at the top (i.e., *M*1), among the first 3 diseases (i.e., *M*3), or among the first 5 diseases (i.e., *M*5) of their differential list. For each checker and doctor, we report the percentages of vignettes that fulfil *M*1, *M*3, and *M*5. The mathematical definitions of *M*1, *M*3, and *M*5 are given in Table 2.

**Table 2:**
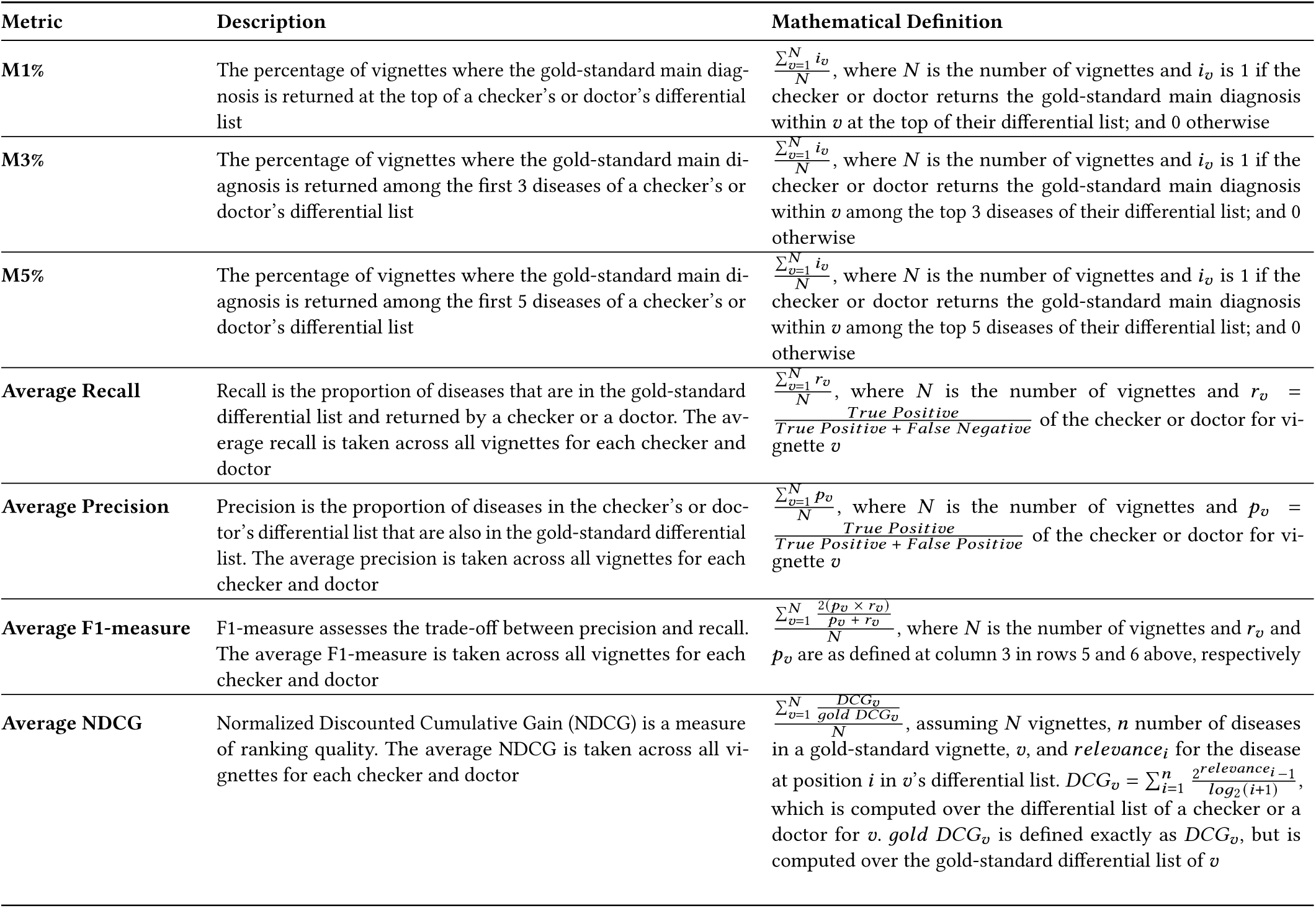
The descriptions and mathematical definitions of the 7 accuracy metrics used in our study.

| Metric | Description | Mathematical Definition |
| --- | --- | --- |
| **M1%** | The percentage of vignettes where the gold-standard main diagnosis is returned at the top of a checker’s or doctor’s differential list | $\frac{\sum_{v=1}^N i_v}{N}$, where $N$ is the number of vignettes and $i_v$ is 1 if the checker or doctor returns the gold-standard main diagnosis within $v$ at the top of their differential list; and 0 otherwise |
| **M3%** | The percentage of vignettes where the gold-standard main diagnosis is returned among the first 3 diseases of a checker’s or doctor’s differential list | $\frac{\sum_{v=1}^N i_v}{N}$, where $N$ is the number of vignettes and $i_v$ is 1 if the checker or doctor returns the gold-standard main diagnosis within $v$ among the top 3 diseases of their differential list; and 0 otherwise |
| **M5%** | The percentage of vignettes where the gold-standard main diagnosis is returned among the first 5 diseases of a checker’s or doctor’s differential list | $\frac{\sum_{v=1}^N i_v}{N}$, where $N$ is the number of vignettes and $i_v$ is 1 if the checker or doctor returns the gold-standard main diagnosis within $v$ among the top 5 diseases of their differential list; and 0 otherwise |
| **Average Recall** | Recall is the proportion of diseases that are in the gold-standard differential list and returned by a checker or a doctor. The average recall is taken across all vignettes for each checker and doctor | $\frac{\sum_{v=1}^N r_v}{N}$, where $N$ is the number of vignettes and $r_v = \frac{\text{True Positive}}{\text{True Positive} + \text{False Negative}}$ of the checker or doctor for vignette $v$ |
| **Average Precision** | Precision is the proportion of diseases in the checker’s or doctor’s differential list that are also in the gold-standard differential list. The average precision is taken across all vignettes for each checker and doctor | $\frac{\sum_{v=1}^N p_v}{N}$, where $N$ is the number of vignettes and $p_v = \frac{\text{True Positive}}{\text{True Positive} + \text{False Positive}}$ of the checker or doctor for vignette $v$ |
| **Average F1-measure** | F1-measure assesses the trade-off between precision and recall. The average F1-measure is taken across all vignettes for each checker and doctor | $\frac{\sum_{v=1}^N \frac{2(p_v \times r_v)}{p_v + r_v}}{N}$, where $N$ is the number of vignettes and $r_v$ and $p_v$ are as defined at column 3 in rows 5 and 6 above, respectively |
| **Average NDCG** | Normalized Discounted Cumulative Gain (NDCG) is a measure of ranking quality. The average NDCG is taken across all vignettes for each checker and doctor | $\frac{\sum_{v=1}^N \frac{DCG_v}{\text{gold } DCG_v}}{N}$, assuming $N$ vignettes, $n$ number of diseases in a gold-standard vignette, $v$, and $relevance_i$ for the disease at position $i$ in $v$’s differential list. $DCG_v = \sum_{i=1}^n \frac{2^{relevance_i - 1}}{\log_2(i+1)}$, which is computed over the differential list of a checker or a doctor for $v$. $\text{gold } DCG_v$ is defined exactly as $DCG_v$, but is computed over the gold-standard differential list of $v$ |

Besides, as in [5, 22, 32], for each tested gold-standard vignette, we use ***recall*** (or *sensitivity* in medical parlance) as a measure of the percentage of relevant diseases that are returned in the checker’s or doctor’s differential list. Moreover, we utilize ***precision*** as a measure of the percentage of diseases in the checker’s or doctor’s differential list that are relevant. For each checker and doctor, we report the average recall and average precision across all vignettes. The average recall and average precision are defined mathematically in Table 2.

Typically, there is a trade-off between recall and precision (the higher the recall, the lower the precision, and vice versa). Thus, in accordance with the standard practice in information retrieval^4^, we further use the ***F1-measure*** that combines the trade-off between recall and precision in one easily interpretable score. The mathematical definition of the F1-measure is provided in Table 2. The higher the F1-measure of a checker or a doctor, the better.

Finally, we measure the ranking qualities of each checker and doctor using the Normalized Discounted Cumulative Gain (***NDCG***) [30] metric that is widely used in practice [63]. To begin with, each disease at position *i* in the differential list of a gold-standard vignette is assigned *relevance*_*i*_. The higher the rank of a disease in the differential list, the higher the relevance of that disease to the correct diagnosis. For instance, if a gold-standard differential list has 3 diseases ordered consecutively as *D*_1_, *D*_2_, and *D*_3_, *relevance*_1_ will be greater than *relevance*_2_, which will be greater than *relevance*_3_. We can assign digits 3, 2, and 1 to *relevance*_1_, *relevance*_2_, and *relevance*_3_, respectively to capture this decreasing relevance from top to bottom in the differential list *D*_1_, *D*_2_, and *D*_3_.

Discounted Cumulative Gain (DCG) is defined mathematically as 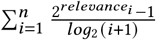, assuming *n* diseases in a vignette’s differential list (see Table 2). As such, DCG penalizes a checker or a doctor if they rank a disease lower in their output differential list than the gold-standard list. For example, if a differential list of a gold-standard vignette, *v*, is *D*_1_, *D*_2_, *D*_3_ and a checker or a doctor produces *D*_3_, *D*_2_, *D*_1_ as a differential for *v*, the DCG of this checker or doctor will be 6.39, while the DCG of *v*’s gold-standard differential is 9.39 (i.e., the checker or doctor was *discounted* 3 points for swapping *D*_3_ with *D*_1_). In contrast, a differential of *D*_1_, *D*_3_, *D*_2_ generated by a checker or a doctor for *v* will result in a DCG of 9.13.

Capitalizing on DCG, Normalized DCG (NDCG) is the ratio of a checker’s or a doctor’s DCG divided by the corresponding gold-standard DCG. Table 2 provides the complete mathematical definition of NDCG. Continuing with the two examples above, if a checker or a doctor outputs *D*_3_, *D*_2_, *D*_1_ as a differential, NDCG would equal 6.39/9.39 = 0.68, while NDCG would equal 9.13/9.39 = 0.97 if a checker or a doctor returns *D*_1_, *D*_3_, *D*_2_. We report the average NDCG across all vignettes for every checker and doctor. To the best of our knowledge, this paper is the first to measure the ranking qualities of the differentials of checkers and doctors.

## 4 RESULTS

### 4.1 Avey versus Checkers

In this section, we present our findings of stage 3. As indicated in Section 3.1, the 400 gold-standard vignettes were tested over 6 checkers, namely, Avey, Ada, WebMD, K Health, Buoy, and Babylon. Not every vignette was successfully diagnosed by every checker. For instance, 18 vignettes failed on K Health because their constituent chief complaints were not available in K Health’s search engine, hence, the sessions could not be initiated. Moreover, 35 vignettes failed on K Health because of an age limitation, whereby only vignettes with ages of 18 years or more were accepted.

Besides search and age limitations, some checkers (in particular, Buoy) crashed while diagnosing certain vignettes, even after trying multiple times. In addition, many checkers did not produce deferential diagnoses for some vignettes albeit concluding the diagnostic sessions. For example, Babylon did not generate deferential diagnoses for 351 vignettes. The reason of why some checkers could not produce diagnoses for some vignettes is uncertain, but we conjecture that it might relate to either not modelling the needed diseases or falling short to recall such diseases despite being modelled. Table 3 summarizes the failure rates and reasons across the examined checkers. Alongside, the table reveals the average number of questions asked by each checker upon successfully diagnosing vignettes.

**Table 3:** Failure reasons and rates as well as success and question counts across the 6 tested checkers (DDx = Differential Diagnosis; Qs = Questions).

|  | Failure Reasons & Counts |  |  | Success Counts |  | Avg. # of Qs |
| --- | --- | --- | --- | --- | --- | --- |
|  | Search Limitations | Age Limitations | Crashed | With No DDx | With DDx |  |
| Avey | 0 | 0 | 0 | 2 | 398 | 24.3 |
| Ada | 0 | 0 | 0 | 0 | 400 | 29.4 |
| WebMD | 2 | 1 | 0 | 3 | 394 | 2.64 |
| K-Health | 18 | 35 | 0 | 2 | 345 | 25.3 |
| Buoy | 2 | 3 | 5 | 74 | 316 | 25.6 |
| Babylon | 15 | 0 | 0 | 351 | 34 | 5.9 |

Figure 3 demonstrates the accuracy results of all the checkers over the 400 vignettes, irrespective of whether they failed or not during some diagnostic sessions^5^. As depicted, Avey outperformed Ada, WebMD, K Health, Buoy, and Babylon by averages of 24.5%, 175.5%, 142.8%, 159.6%, 2968.1% using *M*1; 22.4%, 114.5%, 123.8%, 118.2%, 3392% using *M*3; 18.1%, 79.2%, 116.8%, 125%, 3114.2% using *M*5; 25.2%, 65.6%, 109.4%, 154%, 3545% using recall; 8.7%, 88.9%, 66.4%, 88.9%, 2084% using F1-measure; and 21.2%, 93.4%, 113.3%, 136.4%, 3091.6% using NDCG. Ada was able to surpass Avey by an average of 0.9% using precision, although Avey significantly outpaced it across all the remaining metrics, even with asking an average of 17.2% lesser number of questions (see Table 3). As shown in Figure 3, Avey also outperformed WebMD, K Health, Buoy, and Babylon by averages of 103.2%, 40.9%, 49.6%, 1148.5% using precision, respectively.

**Figure 3:**
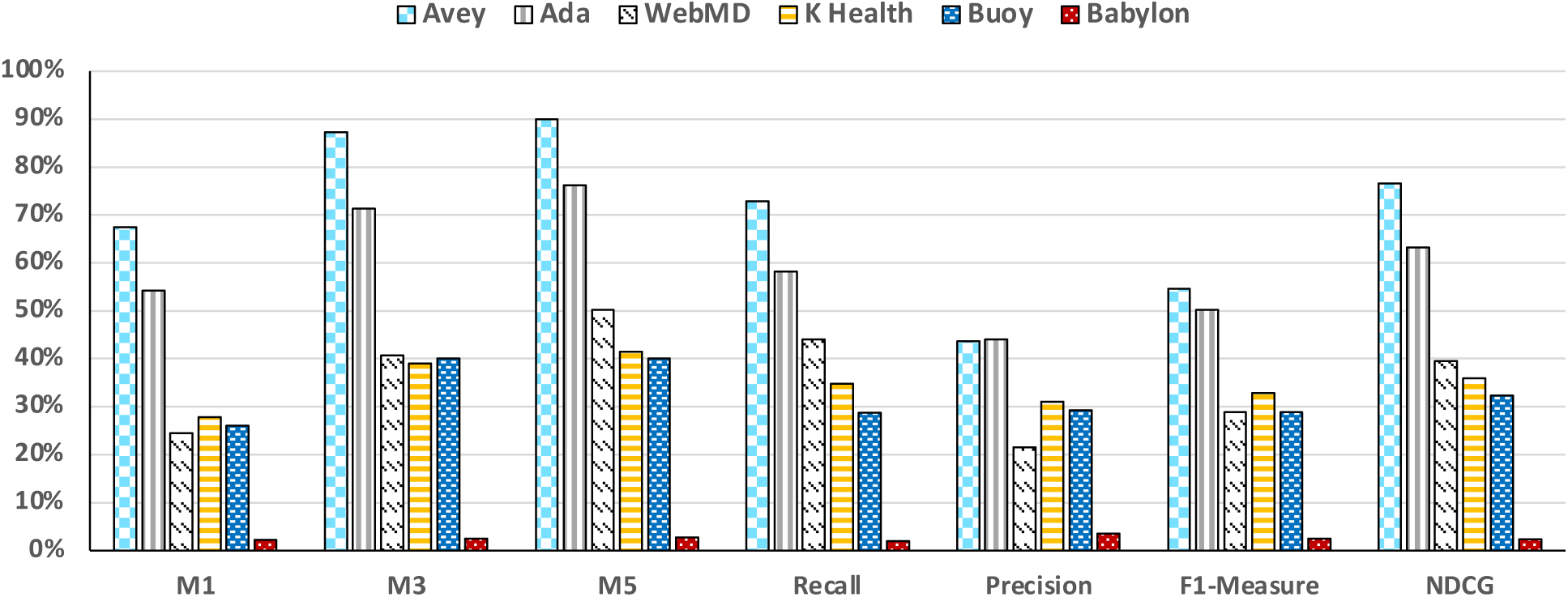
Accuracy results considering for each checker all the succeeded and failed vignettes.

Figure 4 illustrates the accuracy results of all the checkers across only the vignettes that were successful. In other words, checkers were not penalized if they failed to start sessions or crashed during sessions. Nonetheless, Avey still outperformed Ada, WebMD, K Health, Buoy, and Babylon by averages of 24.5%, 173.2%, 110.9%, 152.8%, 2834.7% using *M*1; 22.4%, 112.4%, 94%, 112.9%, 3257.6% using *M*3; 18.1%, 77.8%, 88.2%, 119.5%, 3003.4% using *M*5; 25.2%, 64.5%, 81.8%, 147.1%, 3371.4% using recall; 8.7%, 87.6%, 44.4%, 83.8%, 1922.2% using F1-measure; and 21.2%, 91.9%, 85%, 130.7%, 2964% using NDCG. Under average precision, Ada outpaced Avey by an average of 0.9%, while Avey surpassed WebMD, K Health, Buoy, and Babylon by averages of 101.3%, 22%, 45.6%, 1113.8%, respectively.

**Figure 4:**
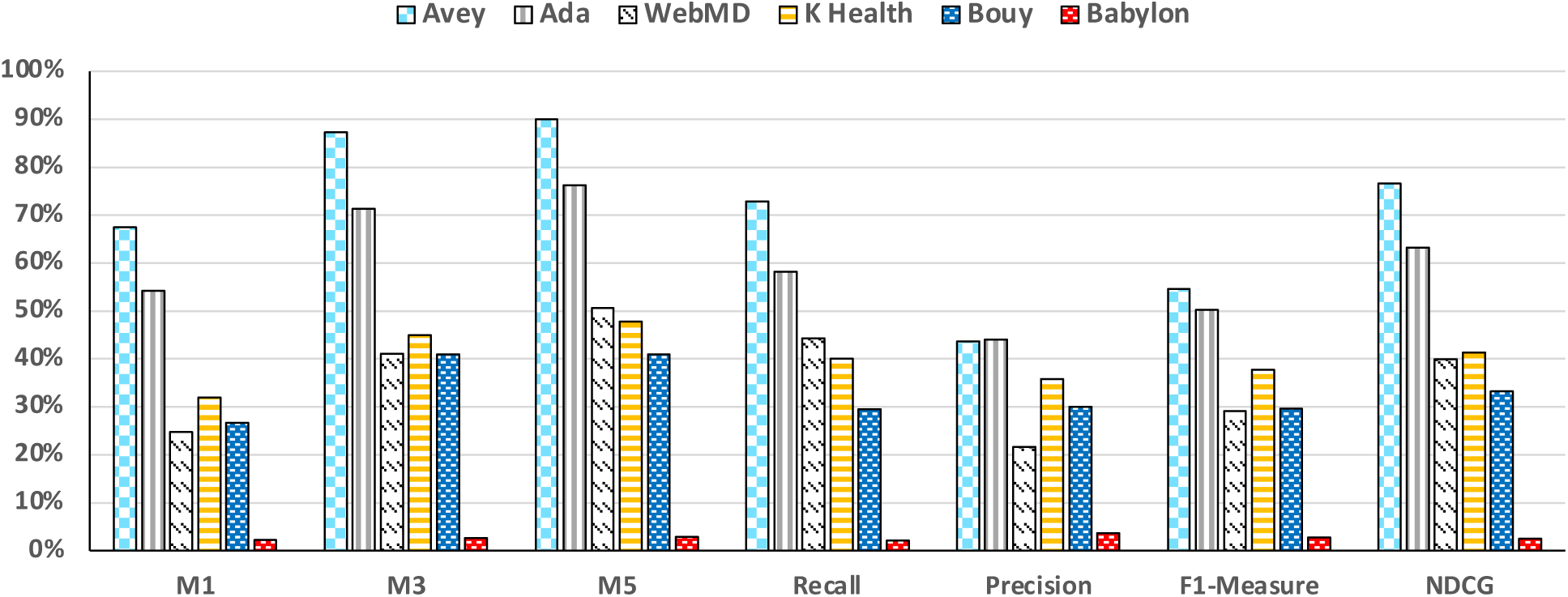
Accuracy results considering for each checker only the succeeded vignettes, with or without differential diagnoses.

Finally, Figure 5 (a) shows the accuracy results of all the checkers over only the vignettes that resulted in differential diagnoses on *every* checker (i.e., the intersection of successful vignettes with differential diagnoses across all checkers). In this set of results, we excluded Babylon since it failed to produce differential diagnoses for 351 out of the 400 vignettes. As demonstrated in the figure, Avey still outperformed Ada, WebMD, K Health, and Buoy by averages of 28.1%, 186.9%, 91.5%, 89.3% using *M*1; 22.4%, 116.3%, 85.6%, 59.2% using *M*3; 18%, 80.1%, 85.7%, 65.5% using *M*5; 23%, 64.9%, 78.5%, 97.1% using recall; 7.2%, 92.7%, 42.2%, 47.1% using F1-measure; and 21%, 93.6%, 77.4%, 76.6% using NDCG. Under average precision, Ada surpassed Avey by an average of 2.4%, while Avey outpaced WebMD, K Health, and Buoy by averages of 109.5%, 20.4%, and 16.9%, respectively.

**Figure 5:**
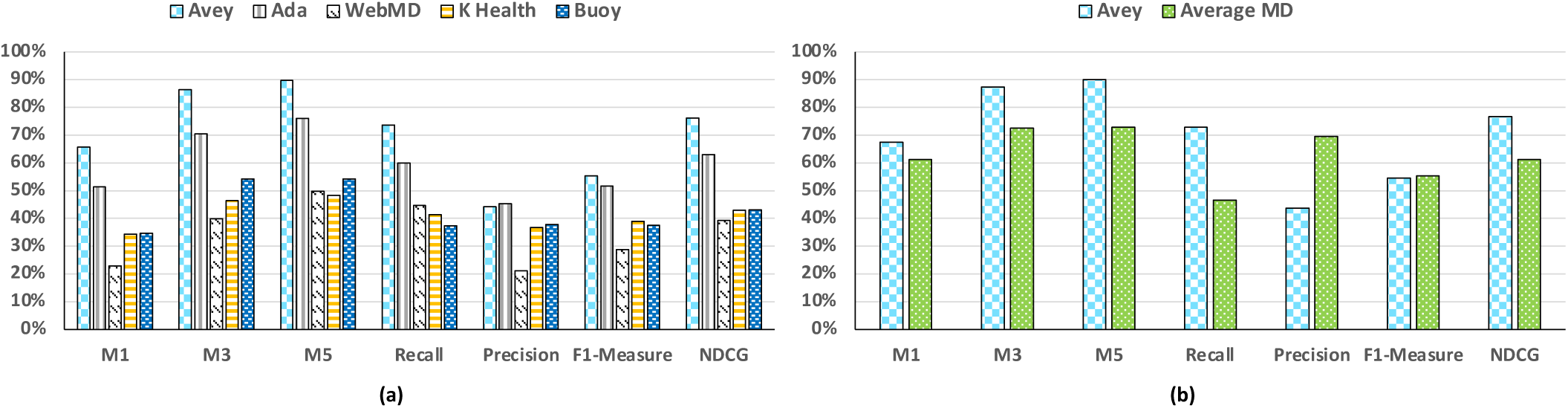
(a) Accuracy results considering only the succeeded vignettes with differential diagnoses across all checkers, and (b) accuracy results of Avey versus three medical doctors, on average (i.e., Average MD).

All the combinations of all the results (i.e., 45 sets of results), including a breakdown between common and less-common diseases, can be found at [50]. In general, Avey demonstrates a superior performance against all the competitor checkers, irrespective of the combination of results.

### 4.2 Avey versus Human Doctors

In this section, we present our findings of stage 4. As discussed in Section 3.1, we tested the 400 gold-standard vignettes on three doctors with an average clinical experience of 16.6 years. Table 4 shows the results of the doctors across all our accuracy metrics. In addition, Figure 5 (b) depicts the results of Avey against *Average MD*, which is the average performance of the three medical doctors. As shown, the human doctors provided average *M*1, *M*3, *M*5, recall, precision, F1-meansure, and NDCG of 61.2%, 72.5%, 72.9%, 46.6%, 69.5%, 55.3%, 61.2%, respectively. In contrast, Avey demonstrated average *M*1, *M*3, *M*5, recall, precision, F1-measure, and NDCG of 67.5%, 87.3%, 90%, 72.9%, 43.7%, 54.6%, 76.6%, respectively.

**Table 4:** Accuracy results of three medical doctors, MD_1_, MD_2_, and MD_3_, with an average experience of 16.6 years.

|  | *M1* | *M3* | *M5* | Recall | Precision | F1-Measure | NDCG |
| --- | --- | --- | --- | --- | --- | --- | --- |
| **MD<sub>1</sub>** | 49.7% | 62% | 62.7% | 41.2% | 58.6% | 48.4% | 52.2% |
| **MD<sub>2</sub>** | 61.3% | 67.2% | 67.5% | 41.2% | 78.1% | 53.9% | 58% |
| **MD<sub>3</sub>** | 72.5% | 88.2% | 88.5% | 57.3% | 71.7% | 63.7% | 73.5% |

To this end, Avey compares favourably to the considered highly experienced doctors, yielding inferior performance in terms of precision and F1-measure, but superior performance in terms of *M*1, *M*3, *M*5, and NDCG. More precisely, the doctors outperformed Avey by averages of 37.1% and 1.2% using precision and F1-measure, while Avey outpaced doctors by averages of 10.2%, 20.4%, 23.4%, 56.4%, and 25.1% using *M*1, *M*3, *M*5, recall, and NDCG, respectively.

## 5 DISCUSSION

### 5.1 Principal Findings

In this paper, we capitalized on the standard clinical vignette approach to evaluate checkers. Specifically, we compiled, peer-reviewed, and utilized 400 vignettes to assess the accuracies of Avey, 5 popular checkers, and 3 primary care physicians with an average experience of 16.6 years. We found that Avey significantly outperforms the 5 checkers and compares favorably to the 3 physicians. To exemplify, under *M*1, Avey outperforms the next best-performing checker, namely, Ada, by 24.5% and the worst-performing checker, namely, Babylon, by 2968.2%. On average, Avey outperforms the 5 checkers by 694.1% using *M*1. In contrast, under *M*1, Avey underperforms the best-performing physician by 6.9% and outperforms the worst-performing one by 35.8%. On average, Avey outperforms the 3 physicians by 13% using *M*1.

In summary, we observed that checkers and physicians vary dramatically in terms of performance. We next order the checkers and physicians (referred to as MD_1_, MD_2_, and MD_3_) from best-performing to worst-performing under each accuracy metric, alongside reporting the statistical ranges and standard deviations.

1. *M*1%: MD_3_, Avey, MD_2_, Ada, MD_1_, K Health, Buoy, WebMD, and Babylon. The range and standard deviation of checkers were 65.3% and 0.21%, while those of physicians were 22.8% and 0.09%, respectively.
2. *M*3%: MD_3_, Avey, Ada, MD_2_, MD_1_, WebMD, Buoy, K Health, and Babylon. The range and standard deviation of checkers were 84.8% and 0.26%, while those of physicians were 26.2% and 0.11%, respectively.
3. *M*5%: Avey, MD_3_, Ada, MD_2_, MD_1_, WebMD, K Health, Buoy, and Babylon. The range and standard deviation of checkers were 87.2% and 0.27%, while those of physicians were 25.8% and 0.11%, respectively.
4. Average Recall: Avey, Ada, MD_3_, WebMD, MD_1_ & MD_2_ (a *tie*), K Health, Buoy, and Babylon. The range and standard deviation of checkers were 70.9% and 0.22%, while those of physicians were 16.1% and 0.08%, respectively.
5. Average Precision: MD_3_, MD_2_, MD_1_, Ada, Avey, K Health, Buoy, WebMD, and Babylon. The range and standard deviation of checkers were 40.6% and 0.13%, while those of physicians were 19.5% and 0.08%, respectively.
6. Average F1-Measure: MD_3_, Avey, MD_2_, Ada, MD_1_, K Health, Buoy & WebMD (a *tie*), and Babylon. The range and standard deviation of checkers were 32.9% and 0.16%, while those of physicians were 15.3% and 0.06%, respectively.
7. Average NDCG: Avey, MD_3_, Ada, MD_2_, MD_1_, WebMD, K Health, Buoy, and Babylon. The range and standard deviation of checkers were 74.2% and 0.23%, while those of physicians were 21.3% and 0.09%, respectively.

### 5.2 Strengths and Limitations of this Study

The strengths and limitations of this study can be summarized as follows:

- The study proposes a comprehensive and rigorous experimentation methodology that taps into the standard clinical vignette approach to evaluate checkers and primary care physicians.
- The study evaluates the performance of checkers and physicians from different angles using 7 standard accuracy metrics, which resulted in the largest set of accuracy metrics that have been considered thus far in the field.
- The study measures for the first time in the field the ranking qualities of the differential diagnoses of checkers and physicians.
- The study presents the largest number of vignettes in the domain so far, which were all peer-reviewed by external and experienced doctors before used for testing checkers and physicians.
- To minimize bias, the checkers were tested by only independent primary care physicians using the peer-reviewed vignettes.
- To facilitate the reproducibility of the study and enable future related studies, all the peer-reviewed vignettes were made publicly and freely available at [49].
- To establish a standard of full transparency and allow for external cross-validation, all the reported results of the checkers and physicians were posted online at [49, 50].
- The physicians that were compared against the checkers may not be a representative sample of primary care physicians.
- The study lacks an evaluation with real patients and covers only 14 body systems with a limited range of conditions.
- The study lacks a comprehensive and rigorous process to choose checkers and considers only a few of them.

### 5.3 Comparison to the Wider Literature

Much work, especially recently, has been done to study checkers from different perspectives. It is not possible to do justice to this large body of work in this short article. As such, we briefly describe some of the most closely related ones, which focus primarily on the accuracy of self-diagnosis.

Semigran *et al*. [53] were the first to study the performance of many checkers across a range of conditions in 2015. They tested 45 vignettes over 23 checkers and discovered that they vary considerably in terms of accuracy, with *M*1 ranging from 5% to 50% and *M*20 (which measures if a checker returns the gold-standard main diagnosis among its top 20 suggested conditions) ranging from 34% to 84%.

Semigran *et al*. published a follow-up paper [52] in 2016 that compared the diagnostic accuracy of physicians against checkers using the same vignettes in [53]. Results showed that, on average, physicians outperformed checkers (72.1% vs 34.0% along *M*1, and 84.3% vs 51.2% along *M*3). However, checkers were more likely to output the gold-standard main diagnosis at the top of their differentials for low-acuity and common vignettes, while physicians were more likely to do it for high-acuity and uncommon vignettes.

The two studies of Semigran *et al*. [52, 53] provided useful insights into the first generation of checkers. However, much has changed since 2015-2016. To exemplify, Gilbert *et al*. [22] recently compiled, peer-reviewed, and tested 200 vignettes over 8 popular checkers and 7 General Practitioners (GPs). As in [53], they found a significant variance in the performance of checkers, but a promise in the accuracy of a new checker, namely, Ada [23]. Ada exhibited accuracies of 49%, 70.5%, and 78% for *M*1, *M*3, and *M*5, respectively. In addition, Ada’s *M*3 was 27.5% higher than that of the next best performing checker (Buoy [25]) and 47% higher than that of the worst-performing one (Your.MD [62]).

None of the checkers in [22] outperformed GPs, but Ada came very close, especially in *M*3 and *M*5. The authors of [22] pointed out that the nature of iterative improvements in software suggests an expected increase in the future performance of checkers, which may at a point in time exceeds that of GPs. As illustrated in Figure 3, we found that Ada is still largely ahead of the conventional checkers, but Avey outperforms it. Furthermore, Avey surpasses physicians under various accuracy metrics as shown in Figure 5 (b).

Hill *et al*. [27] evaluated 36 checkers, 8 of which use AI, over 48 vignettes. They showed that accuracy varies considerably across checkers, ranging from 12% to 61% using *M*1 and from 30% to 81% using *M*10 (where the correct diagnosis appears among the top 10 conditions). They also observed that AI-based checkers outperform rule-based ones (i.e., checkers that do not use AI). Akin to Hill *et al*. [27], Ceney *et al*. [12] detected a significant variation in accuracy across 12 checkers, ranging from 22.2% (CAIDR [11]) to 72% (Ada) using *M*5.

Kannan *et al*. [32] investigated the applicability of learning diagnosis models from electronic health records. They built and presented 3 different machine learning models and showed that they can be effective in generalizing to new patient cases, but with a caveat concerning the number of diseases that they can increasingly incorporate.

Many other studies focused on the diagnostic performance of checkers, but only on a limited set of diagnoses [7–9, 14, 18, 37, 47]. For instance, Berry *et al*. [7] realized that WebMD [58], iTriage [29], and FreeMD [53] are comparable in their performance of delineating between Gastroesophageal reflux disease (GERD) and non-GERD cough. In a follow-up paper [8], they found that 2 out of 3 equally experienced physicians were largely better than WebMD, but not iTriage and FreeMD, on diagnosing patients presenting with cough only. Besides, the third physician was not as good as any of the checkers.

Miller *et al*. [40] presented a real-world usability study of Ada over 523 participants (patients) in a South London primary care clinic over a period of 3 months. Nearly all patients (i.e., 97.8%) found Ada very easy to use. In addition, 22% of patients between ages of 18 and 24 suggested that using Ada before coming to the clinic would have changed their minds in terms of what care to consider next. Studies of other checkers like Buoy and Isabel [28] reported high degrees of utility as well [21, 60].

Some work has also explored the triage capabilities of checkers [5, 51, 60]. Studying the utility and triage capabilities of checkers are beyond the scope of this paper and have been set as future work in Section 5.5.

### 5.4 Implications for Clinicians and Policymakers

As pointed out in Section 1, a UK-based study that engaged 1,071 patients found that more than 70% of individuals between the ages of 18 and 39 years would use a checker [16]. This study was influential in the UK health policy circles, whereby it received press attention and prompted responses from NHS England and NHSX, a UK government policy unit that develops best practices and national policies for technology in health [40, 57]. Given that checkers vary considerably in performance (as shown in Section 4.1), this paper serves in scientifically informing patients, clinicians, and policymakers about the accuracies of some of these checkers.

Besides, this study suggests that a checker should not be publicly launched before it is extensively tested internally and rigorously validated externally. The research, development, and experimental work on Avey took around 4 years and was pursued methodically, thoroughly, and meticulously by a professional team of medical doctors and computer scientists. Avey was launched only after it was verified and tested in-house over thousands of medical cases. Finally, this study suggests that any external scientific validation for any AI algorithm in medicine should be fully transparent and eligible for replication. As a direct translation to this suggestion, we posted all the results of the tested checkers and physicians online as a proof-of-work and to allow for cross-validation. In addition, we made all our peer-reviewed vignettes publicly and freely available. This will not only enable reproducing and validating this study, but further supporting future academic and industry related studies, wherein our gold-standard vignettes can be leveraged as a benchmark suite to test checkers and other similar technologies.

### 5.5 Unanswered Questions and Future Research

This paper focuses solely on studying the diagnostic accuracies of checkers from 7 different standard angles. As such, we set forth 3 immediate and complementary future directions, namely, *usability, utility*, and *extendibility* ones. To elaborate, we will first study the usability and acceptability of Avey with actual patients. In particular, we will investigate how patients perceive Avey and interact with it. During this study, we will observe and identify any barrier in Avey’s UX/UI and language aspects. Afterwards, we will incorporate necessary changes to make Avey’s interface more human-like, especially in terms of ease-of-interaction (e.g., through sound and natural language processing) and friendliness. Second, we will examine how patients respond to Avey’s output and gauge its influence on their subsequent choices for care. Finally, we will extend Avey’s AI model to involve triage and measure its efficacy of referrals and economic impact on patients and healthcare systems.

## 6 CONCLUSIONS

AI-based checkers that undergo rigorous development and testing have the potential to become useful tools for timely, accurate, and instant self-diagnosis. In this paper, we presented Avey, our highly sophisticated and advanced AI-based checker that was extensively researched, designed, developed, and tested for around 4 years before it was launched. We further proposed an experimentation methodology to evaluate Avey against major checkers and seasoned primary care physicians. Results showed that Avey significantly outperforms the considered checkers. In addition, Avey underperforms physicians under some accuracy metrics (e.g., precision and F1-measure), while outperforming them under some others (e.g., *M*1, *M*3, *M*5, recall, and NDCG). In the future, we will extend Avey’s AI model to involve triage and study its usability with real patients and utility for healthcare systems.

## Data Availability

All our gold-standard vignettes are made publicly and freely available at https://avey.ai/research/avey-accurate-ai-algorithm/benchmark-vignette-suite to enable the reproducibility of this work. In addition, all the outputs of the symptom checkers and physicians are posted at the same site to allow for external cross-validation. Lastly, the results of all our 45 sets of experiments are published at https://github.com/rimads/avey-paper/blob/main/main.ipynb to establish a standard of full transparency.

https://github.com/rimads/avey-paper/blob/main/main.ipynb

## Acknowledgements

Vignette review (i.e., vignette standardization, or stage 2 of our experimentation methodology) was carried out by the following independent and experienced physicians: Dr. Zaid Abu Saleh, Dr. Odai Al-Batsh, Dr. Ahmad Alowaidat, Dr. Tamara Altawara, Dr. Arwa Khashan, Dr. Muna Darmach, and Dr. Nour Essale. Vignette testing on checkers (i.e., stage 3 of our experimentation methodology) was carried out by the following independent and experienced physicians: Dr. Maram Alsmairat, Dr. Muna Darmach, and Dr. Ahmad Kakakan. Vignette testing on doctors (i.e., stage 4 of our experimentation methodology) was carried out by the following independent and experienced physicians: Dr. Mohmmad Almadani, Dr. Tala Hamouri, and Dr. Noor Jodeh.

## Contributors

MH conceived the study, designed the experimentation methodology, and supervised the project. SD coordinated the work within and across the project stages (e.g., coordination of vignette creation, vignette standardization, vignette testing on checkers, and vignette testing on doctors). MH conducted the literature review and documentation. SD, MD, and SA created the vignettes and verified the testing results. MD and SS carried out results compilation and summarization. MH and SS carried out data analysis and interpretation. YK developed the web portal for streamlining the processes of reviewing, standardizing, and testing the vignettes. SS maintained Avey’s software and provided technical support. MH wrote the paper. All authors reviewed and commented on drafts of the paper. MH provided administrative support. MH is the guarantor for this work.

## Ethical approval

Not required.

## Patient consent for publication

Not required.

## Data sharing

All our gold-standard vignettes are made publicly and freely available at https://avey.ai/research/avey-accurate-ai-algorithm/benchmark-vignette-suite to enable the reproducibility of this work. In addition, all the outputs of the checkers and physicians are posted at the same site to allow for external cross-validation. Lastly, the results of all our 45 sets of experiments are published at https://github.com/rimads/avey-paper/blob/main/main.ipynb to establish a standard of full transparency.

## Transparency

The guarantor (MH) affirms that the manuscript is an honest, accurate, and transparent account of the study being reported; that no important aspects of the study have been omitted; and that any discrepancies from the study as planned (and, if relevant, registered) have been explained.

This is an Open Access article distributed in accordance with the Creative Commons Attribution Non Commercial (CC BY-NC 4.0) license, which permits others to distribute, remix, adapt, build upon this work non-commercially, and license their derivative works on different terms, provided the original work is properly cited and the use is non-commercial. See: http://creativecommons.org/licenses/by-nc/4.0/.

## Footnotes

1 A finding is defined as a symptom, an etiology, or an attribute, which is a feature of a symptom or an etiology (e.g., in “severe chest pain”, “severe” is an attribute and “chest pain” is a symptom).

2 That is, we always ignore every earlier acceptance and rejection of a vignette if it gets changed at any point in time (no matter how big or small is the change) and start over the reviewing process of the vignette from scratch with all the 7 external doctors.

3 Different checkers and doctors can refer to the same disease differently. As such, our team of physicians considered an output disease by a checker (in stage 3) or a doctor (in stage 4) as a reasonable match to a corresponding disease in the gold standard vignette if the output disease was an alternative name, an umbrella name, or a highly and directly related disease for/to the gold-standard disease.

4 Information retrieval is a field in computer science, wherein the differential diagnosis problem lies partially under.

5 In this set of results, a checker is penalized if it fails to start a session, crashes, or does not produce a differential diagnosis albeit concluding a session.

## REFERENCES

[1] Stephanie Aboueid, Rebecca H Liu, Binyam Negussie Desta, Ashok Chaurasia, and Shanil Ebrahim. 2019. The use of artificially intelligent Self-Diagnosing digital platforms by the general public: Scoping review. JMIR medical informatics 7, 2 (2019), e13445.

[2] Stephanie Aboueid, Samantha Meyer, James R Wallace, Shreya Mahajan, and Ashok Chaurasia. 2021. Young Adults’ Perspectives on the Use of Symptom Checkers for Self-Triage and Self-Diagnosis: Qualitative Study. JMIR Public Health and Surveillance 7, 1 (2021), e22637.

[3] Stephanie Aboueid, Samantha B Meyer, James R Wallace, Shreya Mahajan, Teeyaa Nur, and Ashok Chaurasia. 2021. Use of symptom checkers for COVID-19-related symptoms among university students: a qualitative study. BMJ Innovations 7, 2 (2021).

[4] Saba Akbar, Enrico Coiera, and Farah Magrabi. 2020. Safety concerns with consumer-facing mobile health applications and their consequences: a scoping review. Journal of the American Medical Informatics Association 27, 2 (2020), 330–340.

[5] Adam Baker, Yura Perov, Katherine Middleton, Janie Baxter, Daniel Mullarkey, Davinder Sangar, Mobasher Butt, Arnold DoRosario, and Saurabh Johri. 2020. A comparison of artificial intelligence and human doctors for the purpose of triage and diagnosis. Frontiers in artificial intelligence 3 (2020), 100.

[6] Norman Bates. 2014. Don’t google it. https://vimeo.com/115097884. [Online; accessed 08-Jan-2022].

[7] Andrew C Berry, Nicholas A Berry, Bin Wang, Madhuri Mulekar, Anne Melvin, Richard J Battiola, Frederick K Bulacan, and Bruce B Berry. 2018. Use of online symptom checkers to delineate the ever-elusive GERD versus non-GERD cough. The clinical respiratory journal 12, 12 (2018), 2683.

[8] Andrew C Berry, Nicholas A Berry, Bin Wang, Madhuri S Mulekar, Anne Melvin, Richard J Battiola, Frederick K Bulacan, and Bruce B Berry. 2020. Symptom checkers versus doctors: A prospective, head-to-head comparison for cough. The clinical respiratory journal 14, 4 (2020), 413–415.

[9] Leslie J Bisson, Jorden T Komm, Geoffrey A Bernas, Marc S Fineberg, John M Marzo, Michael A Rauh, Robert J Smolinski, and William M Wind. 2014. Accuracy of a computer-based diagnostic program for ambulatory patients with knee pain. The American journal of sports medicine 42, 10 (2014), 2371–2376.

[10] Shannon Brownlee, Kalipso Chalkidou, Jenny Doust, Adam G Elshaug, Paul Glasziou, Iona Heath, Somil Nagpal, Vikas Saini, Divya Srivastava, Kelsey Chalmers, et al. 2017. Evidence for overuse of medical services around the world. The Lancet 390, 10090 (2017), 156–168.

[11] Caidr. 2006. Symptom Checker. https://caidr.squarespace.com/. [Online; accessed 08-Jan-2022].

[12] Adam Ceney, Stephanie Tolond, Andrzej Glowinski, Ben Marks, Simon Swift, and Tom Palser. 2021. Accuracy of online symptom checkers and the potential impact on service utilisation. Plos one 16, 7 (2021), e0254088.

[13] Christina Cheng and Matthew Dunn. 2015. Health literacy and the Internet: a study on the readability of Australian online health information. Australian and New Zealand journal of public health 39, 4 (2015), 309–314.

[14] Aleksandar Cirkovic. 2020. Evaluation of Four Artificial Intelligence–Assisted Self-Diagnosis Apps on Three Diagnoses: Two-Year Follow-Up Study. Journal of medical Internet research 22, 12 (2020), e18097.

[15] Enrico Coiera, Elske Ammenwerth, Andrew Georgiou, and Farah Magrabi. 2018. Does health informatics have a replication crisis? Journal of the American Medical Informatics Association 25, 8 (2018), 963–968.

[16] Healthwatch Enfield. 2019. Using technology to ease the burden on primary care. https://www.healthwatch.co.uk/reports-library/using-technology-ease-burden-primary-care. [Online; accessed 08-Jan-2022].

[17] United States Medical Licensing Examination. 2019-2020. USMLE Step 2 CK. https://www.usmle.org/step-exams/step-2-ck. [Accessed 05-Feb-2022].

[18] SE Farmer, Matteo Bernardotto, and V Singh. 2011. How good is Internet self-diagnosis of ENT symptoms using Boots WebMD symptom checker? Clinical otolaryngology: official journal of ENT-UK; official journal of Netherlands Society for Oto-Rhino-Laryngology & Cervico-Facial Surgery 36, 5 (2011), 517–518.

[19] Pueyo Ferrer, Martín Baranera, et al. 2017. A new artificial intelligence tool for assessing symptoms in patients seeking emergency department care: the Mediktor application. Emergencias: revista de la Sociedad Espanola de Medicina de Emergencias 29, 6 (2017), 391–396.

[20] John D Firth and Ian Gilmore. 2008. MRCP Part 1 Self-Assessment: Medical Masterclass Questions and Explanatory Answers. Radcliffe Publishing.

[21] Hamish Fraser, Enrico Coiera, and David Wong. 2018. Safety of patient-facing digital symptom checkers. The Lancet 392, 10161 (2018), 2263–2264.

[22] Stephen Gilbert, Alicia Mehl, Adel Baluch, Caoimhe Cawley, Jean Challiner, Hamish Fraser, Elizabeth Millen, Maryam Montazeri, Jan Multmeier, Fiona Pick, et al. 2020. How accurate are digital symptom assessment apps for suggesting conditions and urgency advice? A clinical vignettes comparison to GPs. BMJ open 10, 12 (2020), e040269.

[23] Ada Health. 2011. Symptom Checker. https://ada.com/. [Online; accessed 07-Jan-2022].

[24] Babylon Health. 2013. Symptom Checker. https://www.babylonhealth.com/. [Online; accessed 07-Jan-2022].

[25] Buoy Health. 2014. Symptom Checker. https://www.buoyhealth.com/. [Online; accessed 07-Jan-2022].

[26] K Health. 2016. Symptom Checker. https://khealth.com/. [Online; accessed 07-Jan-2022].

[27] Michella G Hill, Moira Sim, and Brennen Mills. 2020. The quality of diagnosis and triage advice provided by free online symptom checkers and apps in Australia. Medical Journal of Australia 212, 11 (2020), 514–519.

[28] Isabel. 1999. Symptom Checker. https://symptomchecker.isabelhealthcare.com/. [Online; accessed 08-Jan-2022].

[29] iTriage. 2008. Symptom Checker. https://www.aetna.com/. [Not available. Parent organization: Aetna].

[30] Kalervo Järvelin and Jaana Kekäläinen. 2002. Cumulated gain-based evaluation of IR techniques. ACM Transactions on Information Systems (TOIS) 20, 4 (2002), 422–446.

[31] Stefanie Maria Jungmann, Timo Klan, Sebastian Kuhn, and Florian Jungmann. 2019. Accuracy of a Chatbot (ADA) in the diagnosis of mental disorders: comparative case study with lay and expert users. JMIR formative research 3, 4 (2019), e13863.

[32] Anitha Kannan, Jason Alan Fries, Eric Kramer, Jen Jen Chen, Nigam Shah, and Xavier Amatriain. 2020. The accuracy vs. coverage trade-off in patient-facing diagnosis models. AMIA Summits on Translational Science Proceedings 2020 (2020), 298.

[33] Marise J Kasteleyn, Anke Versluis, Petra van Peet, Ulrik Bak Kirk, Jens van Dalfsen, Eline Meijer, Persijn Honkoop, Kendall Ho, Niels H Chavannes, and Esther PWA Talboom-Kamp. 2021. SERIES: eHealth in primary care. Part 5: A critical appraisal of five widely used eHealth applications for primary care– opportunities and challenges. European Journal of General Practice 27, 1 (2021), 248–256.

[34] Doug Knutson. 2018. Family Medicine PreTest Self-Assessment And Review. Mc-Graw Hill Professional.

[35] Sarah Larimer. 2014. Can this ad campaign get people in Belgium to stop Googling their symptoms? https://www.washingtonpost.com/news/to-your-health/wp/2014/11/11/can-this-ad-campaign-get-people-in-belgium-to-stop-googling-their-symptoms/. [Online; accessed 08-Jan-2022].

[36] David M Levine and Ateev Mehrotra. 2021. Assessment of Diagnosis and Triage in Validated Case Vignettes Among Nonphysicians Before and After Internet Search. JAMA network open 4, 3 (2021), e213287–e213287.

[37] Tana M Luger, Thomas K Houston, and Jerry Suls. 2014. Older adult experience of online diagnosis: results from a scenario-based think-aloud protocol. Journal of medical Internet research 16, 1 (2014), e2924.

[38] Seth S Martin, Emmanuel Quaye, Sarah Schultz, Oluwaseun E Fashanu, Jane Wang, Mustapha O Saheed, Prem Ramaswami, Hermes de Freitas, Berthier Ribeiro-Neto, and Kapil Parakh. 2019. A randomized controlled trial of online symptom searching to inform patient generated differential diagnoses. NPJ digital medicine 2, 1 (2019), 1–6.

[39] Ashley ND Meyer, Traber D Giardina, Christiane Spitzmueller, Umber Shahid, Taylor MT Scott, and Hardeep Singh. 2020. Patient Perspectives on the Usefulness of an Artificial Intelligence–Assisted Symptom Checker: Cross-Sectional Survey Study. Journal of medical Internet research 22, 1 (2020), e14679.

[40] Stephen Miller, Stephen Gilbert, Vishaal Virani, and Paul Wicks. 2020. Patients’ utilization and perception of an artificial intelligence–based symptom assessment and advice technology in a British primary care waiting room: exploratory pilot study. JMIR human factors 7, 3 (2020), e19713.

[41] Janet M Morahan-Martin. 2004. How internet users find, evaluate, and use online health information: a cross-cultural review. CyberPsychology & Behavior 7, 5 (2004), 497–510.

[42] Daniel J Morgan, Sanket S Dhruva, Scott M Wright, and Deborah Korenstein. 2016. 2016 update on medical overuse: a systematic review. JAMA internal medicine 176, 11 (2016), 1687–1692.

[43] Government of Australia. 2019. Healthdirect Symptom Checker. https://www.healthdirect.gov.au/symptom-checker. [Online; accessed 08-Jan-2022].

[44] American Board of Family Medicine. 2018. In-Training Examination. https://www.theabfm.org/. [Accessed 05-Feb-2022].

[45] Canadian Institute of Health Information. 2017. Unnecessary Care in Canada. https://www.cihi.ca/sites/default/files/document/choosing-wisely-baseline-report-en-web.pdf. [Online; accessed 08-Jan-2022].

[46] American Academy of Pediatrics. 2020. 2021 PREP Self-Assessment. https://www.aap.org/. [Accessed 05-Feb-2022].

[47] Aimee E Poote, David P French, Jeremy Dale, and John Powell. 2014. A study of automated self-assessment in a primary care student health centre setting. Journal of telemedicine and telecare 20, 3 (2014), 123–127.

[48] CRC Press. 2011-2020. 100 Cases Book Series. https://www.routledge.com/100-Cases/book-series/CRCONEHUNCAS. [Accessed 05-Feb-2022].

[49] Rimads QSTP-LLC. 2022. Avey Benchmark Vignette Suite. https://avey.ai/research/avey-accurate-ai-algorithm/benchmark-vignette-suite. [Accessed 05-Feb-2022].

[50] Rimads QSTP-LLC. 2022. Results of Avey vs. Ada, WebMD, K Health, Buoy, Babylon, and 3 MDs. https://github.com/rimads/avey-paper/blob/main/main.ipynb. [Accessed 05-Feb-2022].

[51] Malte L Schmieding, Rudolf Mörgeli, Maike AL Schmieding, Markus A Feufel, and Felix Balzer. 2021. Benchmarking triage capability of symptom checkers against that of medical laypersons: survey study. Journal of medical Internet research 23, 3 (2021), e24475.

[52] Hannah L Semigran, David M Levine, Shantanu Nundy, and Ateev Mehrotra. 2016. Comparison of physician and computer diagnostic accuracy. JAMA internal medicine 176, 12 (2016), 1860–1861.

[53] Hannah L Semigran, Jeffrey A Linder, Courtney Gidengil, and Ateev Mehrotra. 2015. Evaluation of symptom checkers for self diagnosis and triage: audit study. bmj 351 (2015).

[54] NHS Services. 2019. Babylon GP at hand. https://www.gpathand.nhs.uk/our-nhs-service. [Online; accessed 08-Jan-2022].

[55] Wouter A Spoelman, Tobias N Bonten, Margot WM de Waal, Ton Drenthen, Ivo JM Smeele, Markus MJ Nielen, and Niels H Chavannes. 2016. Effect of an evidence-based website on healthcare usage: an interrupted time-series study. BMJ open 6, 11 (2016), e013166.

[56] Alfred F Tallia, Joseph E Scherger, and Nancy Dickey. 2017. Swanson’s Family Medicine Review E-Book. Elsevier Health Sciences.

[57] Ingrid Torjesen. 2019. Patients find GP online services “cumbersome,” survey finds.

[58] WebMD. 1996. Symptom Checker. https://www.webmd.com/. [Online; accessed 7-Jan-2022].

[59] Ian Wilkinson, Ian Boden Wilkinson, Tim Raine, Kate Wiles, Anna Goodhart, Catriona Hall, and Harriet O’Neill. 2017. Oxford handbook of clinical medicine. Oxford university press.

[60] Aaron N Winn, Melek Somai, Nicole Fergestrom, and Bradley H Crotty. 2019. Association of use of online symptom checkers with patients’ plans for seeking care. JAMA network open 2, 12 (2019), e1918561–e1918561.

[61] Jeremy C Wyatt. 2015. Fifty million people use computerised self triage.

[62] Your.MD. 2013. Symptom Checker. https://www.livehealthily.com/symptom-checker. [Online; accessed 08-Jan-2022].

[63] Xue Zhao. 2013. A Theoretical Analysis of NDCG Ranking Measures. (2013).

